# ^18^F-FAPI PET/CT Imaging in Pneumoconiosis: a new tool for early diagnosis and guiding treatment of pulmonary fibrosis

**DOI:** 10.1101/2025.03.29.25324444

**Authors:** Chaofeng Liu, Zhongyuan Guo, Yiwei Shi, Zhifang Wu, Zhenyu Xiang, Tian Yao, Gang Liang, Zhixing Qin, Ruonan Wang, Li Li, Min Guo, Hailong Wang, Min Pang, Sijin Li

## Abstract

**Purpose:** Pneumoconiosis is characterized by pulmonary fibrosis. The activation of fibroblasts play an important role in the pathological development of pulmonary fibrosis. Chest CT, as a conventional examination to diagnose pulmonary fibrosis of pneumoconiosis, cannot evaluate the fibrosis activity. The application value of ^18^F-FAPI in pneumoconiosis is unclear. This study aimed to clarify the feasibility of ^18^F-FAPI PET/CT in non-invasively monitoring the activity evolution of pulmonary fibrosis in pneumoconiosis and the anti-fibrotic treatment.

**Materials and Methods:** A preliminary clinical study was conducted on 6 pneumoconiosis patients and 4 healthy control individuals, correlation analysis was performed between the ^18^F-FAPI uptake in pulmonary fibrosis areas and the pulmonary diffusing function. Sprague-Dawley rat experiments were performed on three groups concluding pneumoconiosis model, pirfenidone-treated, and normal control groups. ^18^F-FAPI and ^18^F-FDG PET/CT, histopathologic, and hematological analysis were assessed monthly from modeling until 6 months.

**Results:** ^18^F-FAPI uptake in fibrotic areas was found in the pneumoconiosis patients, and negatively correlated with the diffusing function (r = -0.929, *P* = 0.022). In the pneumoconiosis model, ^18^F-FAPI activity preceded one month earlier than relative collagen content (%) in Masson trichrome staining and the level of connective tissue growth factor in plasma, an indicator reflecting the fibroblast activation. The uptake of ^18^F-FAPI, rather than ^18^F-FDG, significantly decreased in the pirfenidone-treated group compared to the pneumoconiosis group (*P* < 0.05).

**Conclusion:** ^18^F-FAPI PET/CT imaging holds promise for the early identification of pulmonary fibrosis activity and monitoring its evolution in pneumoconiosis, offering a precise clinical opportunity for targeted anti-fibrotic treatment.

## Introduction

Pneumoconiosis is a pulmonary interstitial fibrotic disease characterized by structural lung tissue damage and impaired lung function. This condition arises from prolonged inhalation of mineral dust particles with varying pathogenicity(1). Recent studies increasingly indicate a link between pulmonary fibrosis and Fibroblast Activation Protein (FAP). The pulmonary inflammation and fibrosis associated with pneumoconiosis can be commonly observed via High-Resolution Computed Tomography (HRCT) in clinical practices(2). Unfortunately, HRCT cannot distinguish the early or late stage of fibrotic lesions and their activities(3, 4), which may lead to repeated tests, misdiagnosis, and unnecessary medical care(5). Therefore, there is an urgent need for a sensitive and non-invasive method to detect and diagnose active fibrotic lesions earlier than HRCT in pneumoconiosis, and to monitor therapy responses.

FAP-specific inhibitors (FAPIs) have been developed into radiopharmaceuticals and applied to *in vivo* PET/CT (Positron Emission Tomography/Computed Tomography) imaging of activated fibroblasts, such as malignant tumors(6), and systemic sclerosis-associated interstitial lung disease (ILD)(7). However, it remains unclear whether ^18^F-FAPI PET/CT imaging is feasible in non-invasively visualizing the activity and progression of pulmonary fibrosis lesions, and further, detecting the efficacy of anti-fibrotic treatment.

In this study, we preliminarily observed ^18^F-FAPI uptake in pneumoconiosis patients and analyzed the correlation with pulmonary functional parameters. Further, an in-vivo and ex-vivo animal experiment concluding pneumoconiosis model, pirfenidone-treated, and normal control groups were studied, compared with commonly used ^18^F-FDG inflammation imaging.

## Materials and methods

### Clinical study

#### Participants enrollment

This single-center prospective study was approved by the Clinical Research Ethics Committee of the First Hospital of Shanxi Medical University (KYLL-2023-130). All participants signed the informed consent form.

We confirmed patients with pneumoconiosis by diagnostic-level chest CT, aged 18-80 years, between Oct 1, 2022, and Sep 30, 2023. Exclusion criteria were as follows: patients with mild pulmonary fibrosis in HRCT, tuberculosis, malignancy, refusal to participate, prior lung surgery, pulmonary hypertension, inability to give informed consent, and sarcoidosis. All assessments were done by the senior chief physician in the Interstitial Lung Disease Subspecialty Group, based on the medical histories, clinical data, and auxiliary laboratory examinations. Healthy individuals without lung diseases were confirmed by chest CT as controls. All confirmatory chest CTs were conducted in one month before inclusion.

#### Pulmonary function test for participants

All enrolled subjects underwent pulmonary function tests within 1 week before PET/CT imaging (Discovery VCT, General Electric Company). The parameters included the forced vital capacity (FVC) for the ventilation function and the carbon monoxide diffusion capacity (DL_CO_) for the diffusion function.

#### 18F-FAPI PET/CT imaging and analysis

The participants performed ^18^F-FAPI PET/CT imaging without any preparation. Static PET/CT scans (2 min/bed, 5-7 beds) were performed 10 min, 60 min, and 180 min after the ^18^F-FAPI (244.2-358.9 MBq) injection intravenously.

Images were transferred to and analyzed on the Advanced Workstation (Version 47, GE Healthcare) after acquisition and reconstruction. PET/CT fusion images in the coronal, axial, and sagittal planes were reviewed. The outlining and measurement of the volume of interest (VOI) were done separately by two experienced senior nuclear medicine physicians, the average values were taken. The bilateral lungs were divided into 10 equal levels from the lung apex to the lung base, and on each level, the VOI was manually outlined 0.5 cm from the outer surface. The software automatically generated the SUVmean, and the mean value of all the levels was calculated as the whole lung SUVmean. On the CT image, fibrotic and nonfibrotic areas were distinguished, and two spheres with 1 cm^3^ were placed respectively to obtain the SUVmean. Muscle tissue was used as background, and spheres with 1 cm^3^ were placed to obtain the SUVmean. The ratio of SUVmean of the lung and the muscle was used to determine the target-to-background ratio (TBRmean).

### Animal experiments

#### *In vivo* pneumoconiosis modeling

All animal experiments were approved by the Animal Ethics Committee and the Animal Experimentation Control and Supervision Committee of Shanxi Medical University. Male SD rats (aged 6-8 weeks) were purchased from the Laboratory Animal Center of Shanxi Medical University. All rats were housed in pathogen-free micro isolator cages under controlled light, temperature, and humidity at the Collaborative Innovation Center for Molecular Imaging. The rats were fed commercial chow (SPF, Beijing Biotechnology Co., Ltd.) and purified water *ad libitum*. All rats experienced one week of acclimatization in the facility before the experiment started.

All the rats were randomly divided into three groups: the control, pneumoconiosis, and pirfenidone therapy group (6 rats/group). Each rat was anesthetized with isoflurane (VETEASY, R510-22-10, RWD Life Science Co., Ltd) and weighed. The pneumoconiosis model was established by administration of SiO_2_ (0.5 mL of 50 mg/mL, 1-5 μm silica particles, s5631, Sigma-Aldrich) in single tracheal dosing. For the control group, 0.5 mL saline was injected instead. A Micro CT scan (Inviscan Imaging Systems, France) was performed 7 days after intratracheal administration to verify the pneumoconiosis model. The pirfenidone therapy group was administered with Pirfenidone (10 mg/kg, ETUARY, Beijing Continent Pharmaceuticals Co., Ltd.) via the esophagus using a gavage needle (55mm, 16G) with a 2.5 mm ball tip once a day for 6 months from Day 7 after modeling verification. *In vivo* PET/CT imaging was performed each month thereafter.

#### *In vivo* imaging of active fibrosis and inflammation by ^18^F-FAPI and ^18^F-FDG PET/CT

Synthesis and labeling of ^18^F-FAPI was performed as described previously(8). Rats were injected with 6-8 MBq of tracer in a volume of 500-700 μL via the tail vein. Micro PET/CT (Inviscan Small Animal *In Vivo* Imaging System, France) imaging was conducted 60 min after injection, the acquisition time was 10 min. PET images were reconstructed via the 3D-ordered-subsets implementations-Monte Carlo algorithm. For ^18^F-FDG PET/CT imaging, rats were fasted overnight the day before.

PMOD software (Version 4.1) was used for image analysis. The bilateral lungs were manually outlined at each slice from the apices to the base on CT trans-axial images, with 0.5 cm apart from the lung margins. The mean standardized uptake value (SUVmean) of whole-lung was automatically generated to reflect the level of the tracer uptake. The SUVmean of the lung tissue/SUVmean of the muscle tissue was used to determine the target-to-background ratio (TBRmean).

### Histology examination

The lung was harvested for histological analysis. Hematoxylin and eosin (HE) and Masson trichrome staining were used to detect lung inflammation and fibrosis. FAP expression levels and the percentages of FAP+ areas were tested using immunofluorescence (IF) staining and Immunohistochemistry (IHC) in the lung tissue of pneumoconiosis rats. The details were available in the Supplemental methods S1-S3.

### Real-time Quantitative PCR (qPCR)

FAP mRNA expression was tested using qPCR in the lung of pneumoconiosis rats (Supplemental method S4).

### Enzyme-linked immunosorbent assay (ELISA)

The inflammatory cytokines and fibrogenic cytokines of plasma in pneumoconiosis rats were measured by ELISA (Supplemental method S5).

### Statistical analysis

SPSS software (Version 26.0) was used for statistical analysis. Data were expressed as the mean value

± standard deviation. Comparisons were performed using independent sample t-tests and ANOVA. Pearson correlation was used to evaluate the associations between SUVmean and DL_CO_%, and the associations ^18^F-FAPI uptake between fibrotic and nonfibrotic areas in pneumoconiosis patients. A two-tailed *P*-value of less than 0.05 was considered significant.

## Results

### Clinical Study Results

#### Baseline Characteristics

4 healthy control individuals (50.50 ± 13.77 years old) and 6 pneumoconiosis patients (62.50 ± 5.57 years old) were recruited and underwent the full study procedure (Figure. 1). The baseline characteristics and pulmonary function of the participants were shown in Table 1. The predicted FVC% (91.7 ± 16.21% vs 95.17 ± 1.57%) and DL_CO_% (70.58 ± 16.37% vs 84.46 ± 3.66%) of the pneumoconiosis patients were lower than those in the control individuals (*P* > 0.05), indicating reduced diffusion function.

**FIGURE 1.**
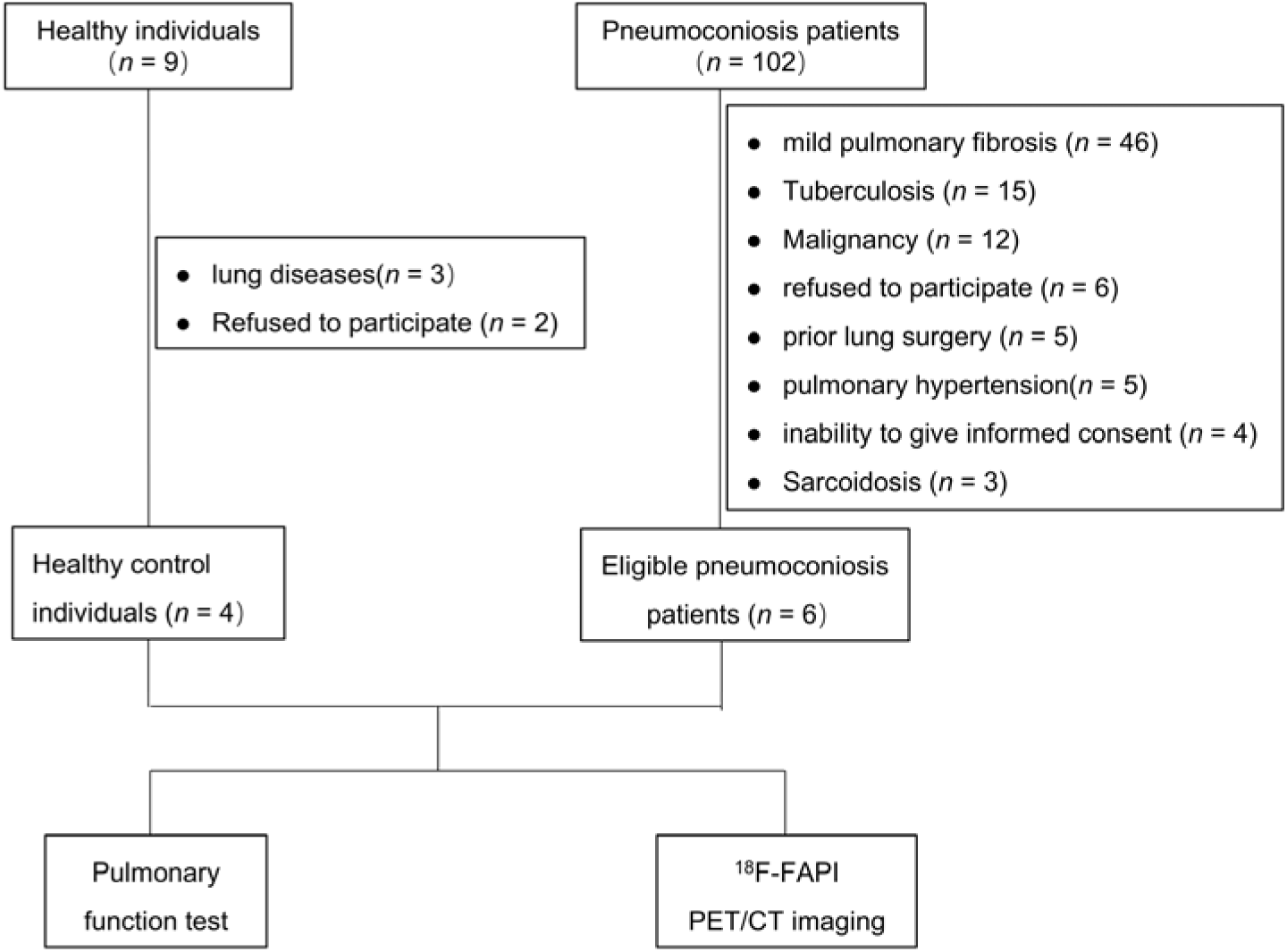
Enrollment and design of the clinical study

**TABLE 1.**
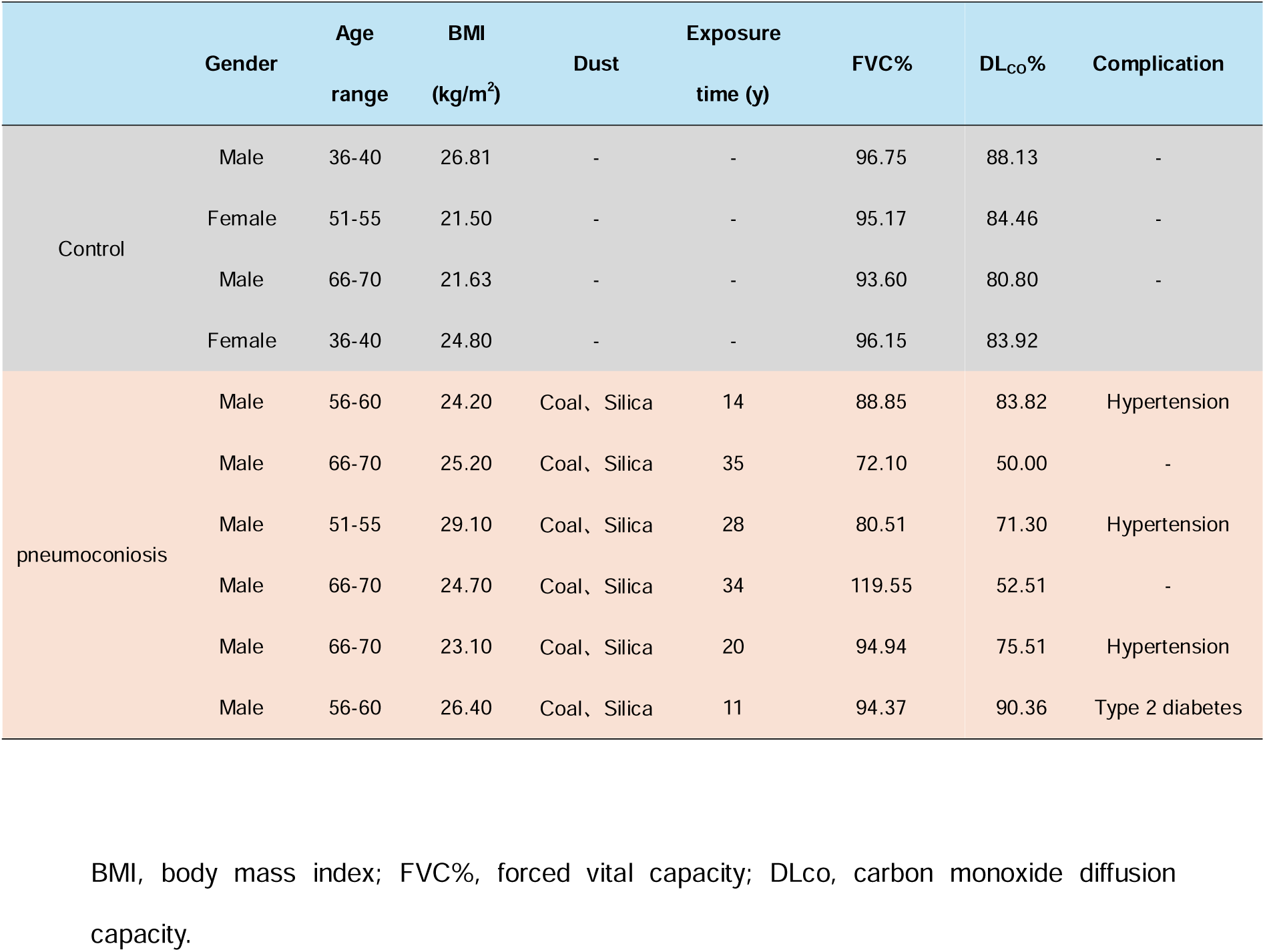
Baseline Characteristics of Healthy control individuals and pneumoconiosis.

|  | Gender | Age<br>range | BMI<br>(kg/m<sup>2</sup>) | Dust | Exposure<br>time (y) | FVC% | DL<sub>co</sub>% | Complication |
| --- | --- | --- | --- | --- | --- | --- | --- | --- |
| Control | Male | 36-40 | 26.81 | - | - | 96.75 | 88.13 | - |
|  | Female | 51-55 | 21.50 | - | - | 95.17 | 84.46 | - |
|  | Male | 66-70 | 21.63 | - | - | 93.60 | 80.80 | - |
|  | Female | 36-40 | 24.80 | - | - | 96.15 | 83.92 | - |
| pneumoconiosis | Male | 56-60 | 24.20 | Coal, Silica | 14 | 88.85 | 83.82 | Hypertension |
|  | Male | 66-70 | 25.20 | Coal, Silica | 35 | 72.10 | 50.00 | - |
|  | Male | 51-55 | 29.10 | Coal, Silica | 28 | 80.51 | 71.30 | Hypertension |
|  | Male | 66-70 | 24.70 | Coal, Silica | 34 | 119.55 | 52.51 | - |
|  | Male | 66-70 | 23.10 | Coal, Silica | 20 | 94.94 | 75.51 | Hypertension |
|  | Male | 56-60 | 26.40 | Coal, Silica | 11 | 94.37 | 90.36 | Type 2 diabetes |

BMI, body mass index; FVC%, forced vital capacity; DL<sub>co</sub>, carbon monoxide diffusion capacity.

The typical PET/CT images of the pneumoconiosis patients and the health control individuals were displayed in Figure. 2A. Healthy controls exhibited slight ^18^F-FAPI uptake in the whole lung, whereas pronounced uptake was observed under the pleura of pneumoconiosis patients. The SUVmean of ^18^F-FAPI in the whole lungs peaked at 10 min after injection in both two groups and then decreased with time. Notably, the greatest uptake difference between the groups was observed at 60 min (*P* < 0.01), as shown in Figure. 2B.

**FIGURE 2.**
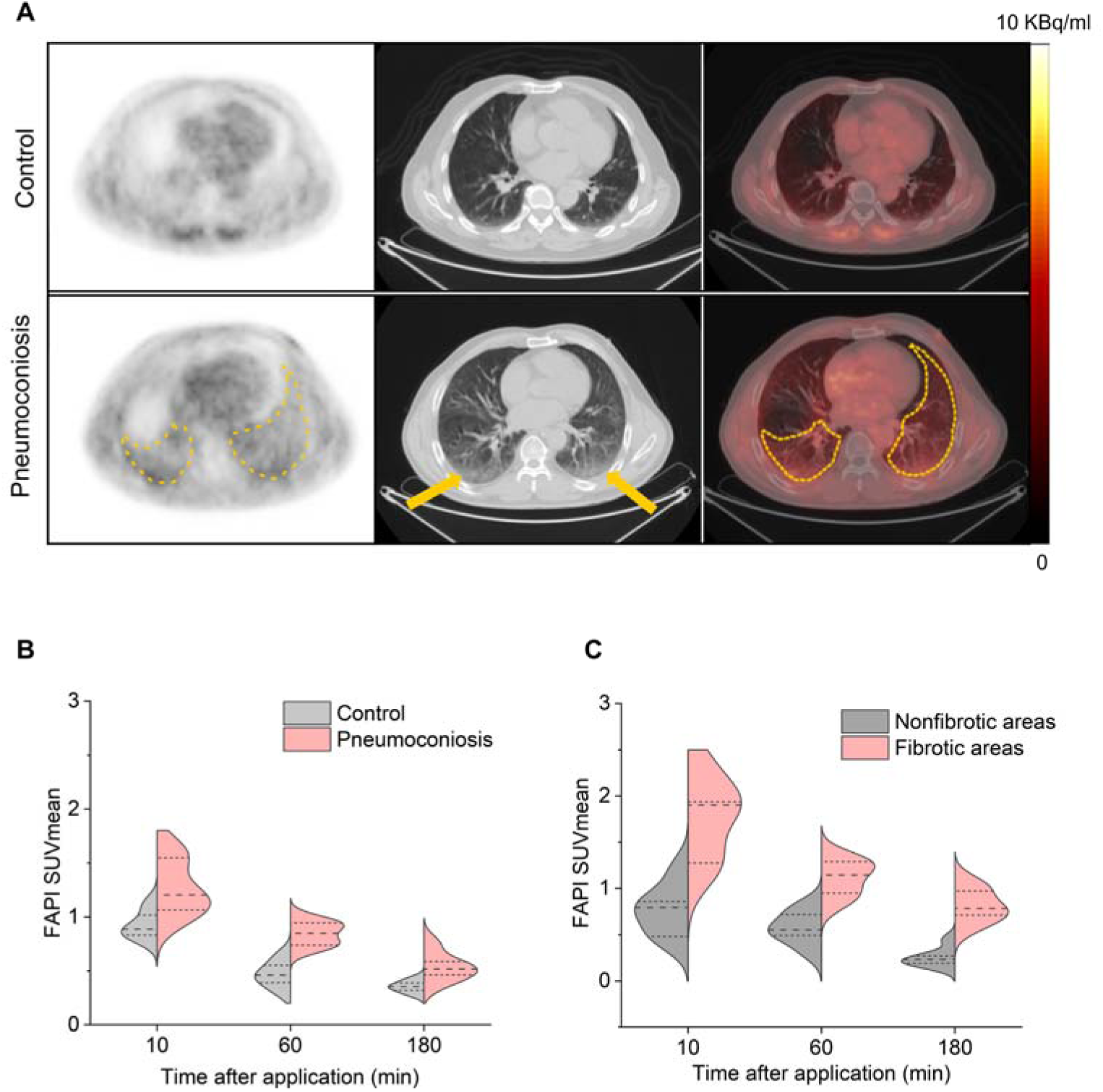
FAP targeted PET/ CT imaging of healthy control individuals and pneumoconiosis patients. (A) The columns from left to right successively showed representative images of ^18^F-FAPI uptake in lung cross-section with PET, CT, and PET/CT. The yellow dashed boxes represent the active fibrosis, and the yellow arrows represent fibrosis. (B) SUVmean of ^18^F-FAPI in healthy control individuals and pneumoconiosis patients at 10 min, 60 min, and 180 min after injection. (C) SUVmean of ^18^F-FAPI in fibrotic and nonfibrotic areas of pneumoconiosis patients at 10 min, 60 min, and 180 min after injection. FAP, Fibroblast activation protein; PET/CT, positron emission tomography/computed tomography.

#### Comparison of ^18^F-FAPI uptake between fibrotic and nonfibrotic areas in pneumoconiosis patients

The SUVmean of ^18^F-FAPI in the fibrotic areas of pneumoconiosis patients peaked 10 minutes post-injection and then gradually declined, which was significantly higher in the fibrotic regions of pneumoconiosis compared to the nonfibrotic regions (*P* < 0.05, Figure. 2C).

#### Correlation analysis between SUVmean and DL_CO_%

The SUVmean in fibrotic lung regions exhibited a significant negative correlation with DL_CO_% (r = -0.929, *P* = 0.022). No statistically significant associations were observed between FVC% and tracer uptake either in non-fibrotic areas or across the entire lung.

### Animal experiments results

#### Active fibrosis *in vivo*pneumoconiosis modeling

The ^18^F-FAPI uptake in the pneumoconiosis group increased to a peak in the Month 4, and then decreased, which was higher than that in the pirfenidone therapy group (Figure. 3A). The TBRmean of the ^18^F-FAPI in the pneumoconiosis group in the Month 4 (3.85 ± 0.58), and the Month 5 (3.41 ± 0.48) were significantly higher than that before modeling (2.13 ± 0.17), and the TBRmean of the pirfenidone therapy group was statistically lower than that in the pneumoconiosis group in the Month 4 (2.93 ± 0.26 vs. 3.85 ± 0.58), and the month 5 (2.55 ± 0.20 vs. 3.41 ± 0.48) (*P* < 0.05) (Figure. 3C). Masson trichrome staining showed gradually increased collagen fiber deposition and fibrosis in the pneumoconiosis group, and relative collagen content (%) increased in the Month 5, and 6 compared to the other timepoints (*P*<0.001). The pirfenidone therapy group had less fibrosis than the pneumoconiosis group (Figure. 3B and Figure. 3D). FAP immunofluorescence staining showed the expression of FAP in the pneumoconiosis group peaked in the Month 4, and decreased in the Month 5 (Figure. 4A and Figure. 4B). The FAP concentration of the pneumoconiosis group increased in the Month 1, 4, and 6 compared to the Month 0, with the highest concentration in the Month 4 and significant differences compared to the Month 0, 1 and 6 (all *P* < 0.001, Figure. 4D and Figure. 4E). In addition, the FAP mRNA in the pneumoconiosis group showed the same variation trends as the FAP concentration (Figure. 4C).

**FIGURE 3.**
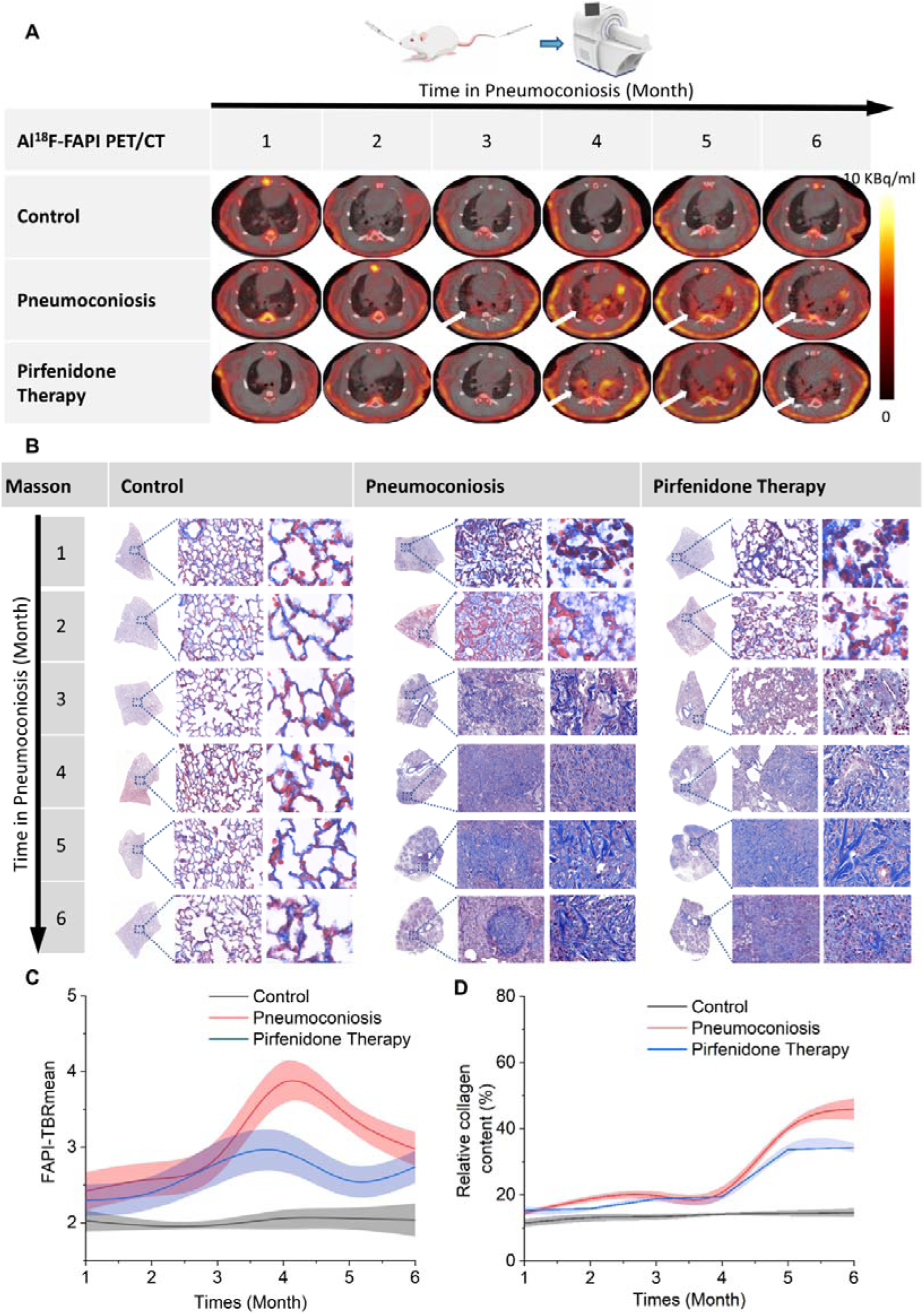
Evaluation of pulmonary fibrosis processes. (A) A series of sequential ^18^F-FAPI PET/CT images of control, pneumoconiosis, and pirfenidone-therapy rats in the month 1, 2, 3, 4, 5, and 6. The white arrows represent the active fibrosis. (B) Masson trichrome staining of lung tissue of the control, pneumoconiosis, and pirfenidone-therapy rats in different magnifications (Macrograph, ×100, ×400) in the month 1, 2, 3, 4, 5, and 6. (C) Quantitative analysis of the ^18^F-FAPI TBRmean in the control, pneumoconiosis, and pirfenidone-therapy rats in the month 1, 2, 3, 4, 5, and 6. (D) Relative collagen content (%) in Masson trichrome staining of the lung of the control, pneumoconiosis, and pirfenidone-therapy rats in the month 1, 2, 3, 4, 5, and 6. TBR, target-to-background ratio.

**FIGURE 4.**
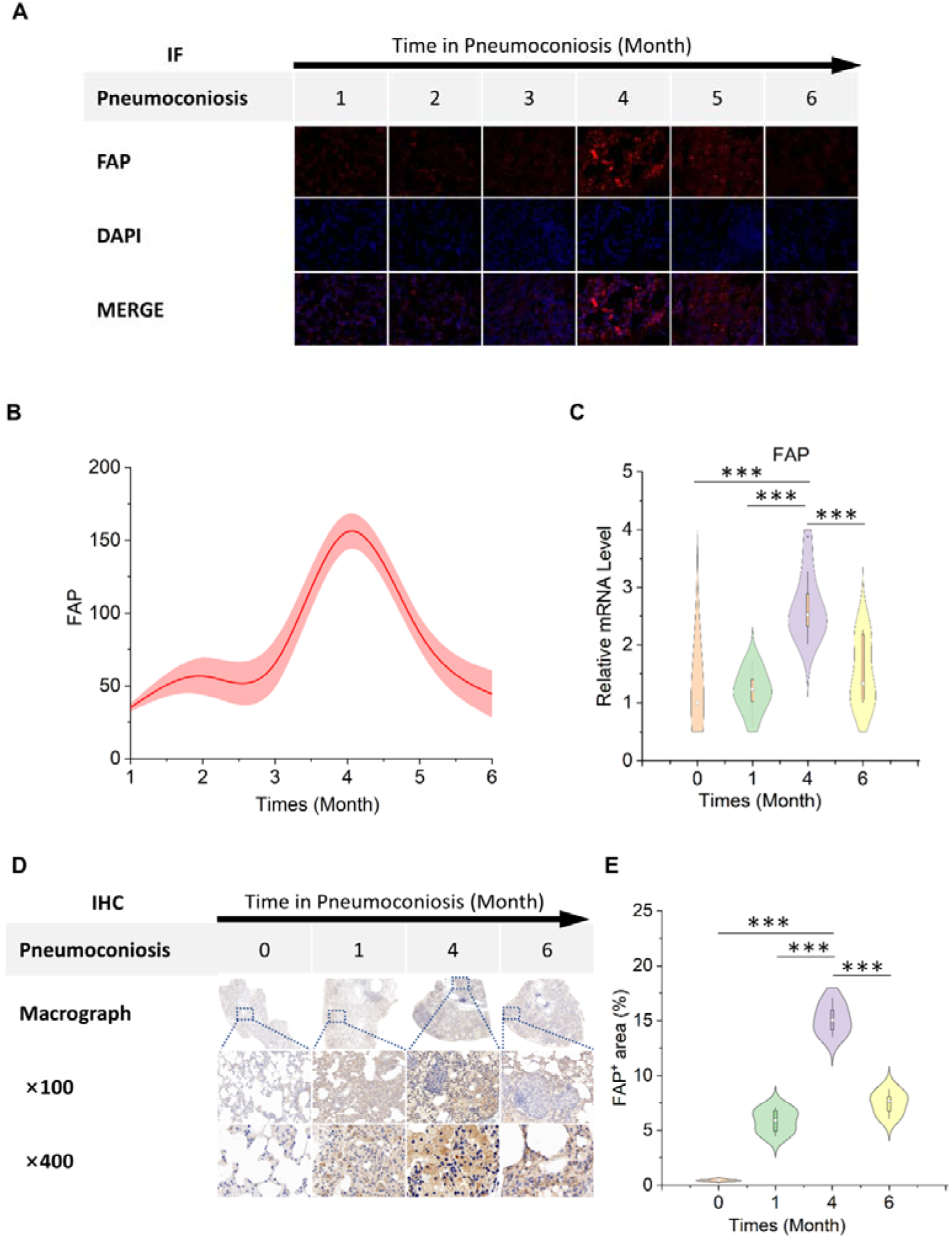
The expression of FAP in pneumoconiosis rats. (A) Representative FAP IF staining in the Month 1, 2, 3, 4, 5, 6. (FAP, red; DAPI, blue; Scale bar: 100 μm). (B) The FAP levels by immunofluorescence staining in the lung. (C) The FAP mRNA expression of rat lung in the Month 0, 1, 4, 6(n = 4, p < 0.001). (D) Representative FAP IHC staining in the Month 0, 1, 4, and 6 in different magnifications (Macrograph, ×100, ×400). (E) The percentages of FAP^+^ areas in Immunohistochemical staining sections of lung tissue in the Month 0, 1, 4, and 6 (n = 4, p < 0.001). FAP, fibroblast activation protein; DAPI, 4’,6-diamidino-2-phenylindole; IF, immunofluorescence; IHC, immunohistochemical.

#### The evolution of inflammation after pneumoconiosis modeling

The ^18^F-FDG uptake of rat lungs in the pneumoconiosis was increased after modeling, peaked in the Month 3, and decreased gradually. The ^18^F-FDG uptake in the pirfenidone therapy group was lower than that in the pneumoconiosis group in the Month 3-6 (Figure. 5A). The SUVmean displayed no significant difference between the pirfenidone therapy group and the pneumoconiosis group (*P* > 0.05, Figure. 5C). HE staining images showed that the degree of inflammation gradually increased in the pneumoconiosis group, with multiple granulomas forming in the later stage (Figure. 5B). IL-6, TNF-L, and CTGF levels all increased after establishing models, started to increase significantly respectively in the Month 2, 4, and 5 compared to the Month 0 (*P* < 0.05, Figure. S1A; *P* < 0.001, Figure. S1B; *P* < 0.001, Figure. S1C).

**FIGURE 5.**
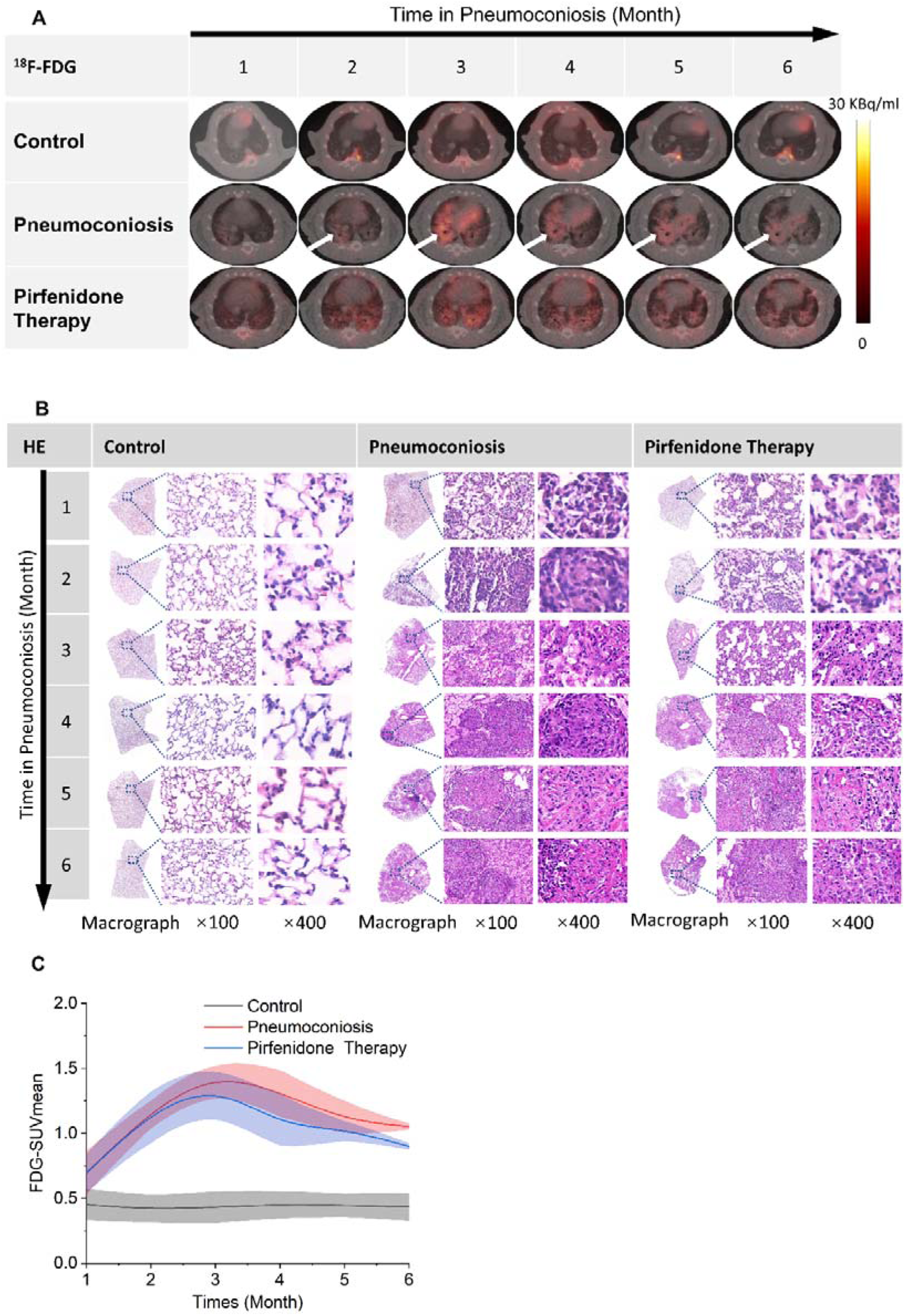
Evaluation of pulmonary inflammation processes. (A) A series of sequential ^18^F-FDG PET/CT images of control, pneumoconiosis, and pirfenidone-therapy rats in the month 1, 2, 3, 4, 5, and 6. The white arrows represent the active inflammation. (B) HE staining of lung tissue sections of the control, pneumoconiosis, and pirfenidone-therapy rats in different magnifications (Macrograph, ×100, ×400) in each month. (C) Quantitative analysis of the ^18^F-FDG uptake (SUVmean) in the lung of the control, pneumoconiosis, and pirfenidone-therapy rats. SUV, standardized uptake value.

## Discussion

In this study, ^18^F-FAPI PET/CT was employed as a pivotal radioactive tracer for visualizing lung fibroblasts and meticulously monitoring the progression of pulmonary fibrosis in pneumoconiosis. Originally a significant accumulation of ^18^F-FAPI within the regions of the fibrotic lung was observed in pneumoconiosis patients, an occurrence inversely correlated with pulmonary diffusion capacity - a harbinger of adverse clinical prognosis. Extending these observations to the rat model of pneumoconiosis, this study unveiled that ^18^F-FAPI accumulated in lung tissue and achieved its zenith post the inaugural administration of silica, thereafter, exhibiting a decline commensurate with the development of pulmonary fibrosis. The uptake patterns of ^18^F-FAPI exhibited a commendable concordance with the FAP IF staining outcomes, but preceded the fibrotic manifestations delineated by Masson’s trichrome staining. These preliminary findings collectively suggest that ^18^F-FAPI PET/CT imaging holds promise as a frontline diagnostic modality capable of identifying patients in the active fibrotic phase of pneumoconiosis at an early stage, thereby offering prognostic insights crucial for informing therapeutic strategies. This dual capability of early detection and prognostication underscores the potential of ^18^F-FAPI PET/CT as a transformative tool in the treatment decisions of pneumoconiosis.

Pulmonary fibrosis stands as a paramount clinical determinant of pneumoconiosis progression; its pathogenesis, shrouded in mystery, remains elusive. Within this intricate web, the activation of fibroblasts and the abnormal deposition of extracellular matrix weave a dual tapestry of destruction. This process not only fosters the formation of lung fibrotic scars but also relentlessly advances toward respiratory failure(9, 10). FAP, a biomarker signifying activated fibroblasts, manifests its elevated expression in pulmonary fibrosis tissue as a harbinger of impending fibroblast foci development(11–13). Detecting these processes and quantitatively analyzing the expression levels of FAP should provide a scientific basis for the early prevention and treatment of pneumoconiosis and pulmonary fibrosis. Qin *et al*. detected the dynamic in lung injury using a rat model of radiation-induced lung injury, specifically employing ^18^F-FAPI PET/CT imaging. Different from our research findings, ^18^F-FAPI uptake decreased to baseline levels in weeks 2-3 after irradiation, with a secondary increase in week 4 and stabilization during weeks 4-6(14). The potential explanation is that radiation-induced lung injury leads to radiation pneumonia in the early stages and progresses to radiation-induced lung fibrosis in later stages. Conversely, in pneumoconiosis, inflammation persists throughout the entire course, with pulmonary fibrosis manifesting only in the advanced stages, whose progression has been corroborated by ^18^F-FDG imaging. Yang *et al*. compared lung tissues from silicosis patient post-transplantation with those of healthy organ donors. Their findings revealed that 12 out of the studied silicosis patients exhibited elevated levels of FAP protein compared to healthy donors, a result consistent with our study(4).

^18^F-FDG PET/CT imaging was employed to investigate the correlation between inflammation and fibrosis and to evaluate the effectiveness of anti-fibrotic therapy. Our findings indicate persistent active inflammation in fibrotic lesions from the onset through the progression of pneumoconiosis, accompanied by a suboptimal therapeutic response to pirfenidone. This implies that ^18^F-FDG has limited utility in assessing active fibrosis in pneumoconiosis(15). The uptake of ^18^F-FDG reflects the recruitment of neutrophils, macrophages, and fibroblasts involved in inflammatory and fibrotic processes during the development of pneumoconiosis. Kubota *et al*. reported that a significant component of ^18^F-FDG uptake by tumor tissues originates from active inflammatory cells surrounding the tumor, which are more likely to take up ^18^F-FDG than tumor cells themselves(16). The interplay of multiple factors in the inflammatory course can continuously increase FDG uptake in active inflammatory disorders. PET/CT imaging can delineate the location, extent, and severity of inflammatory lesion involvement(17, 18). However, it remains challenging to discern which pathological alterations predominate at any given stage, thereby complicating the selection of optimal timing and therapeutic regimen. A clinical investigation by Bondue *et al*. revealed no significant changes in ^18^F-FDG uptake three months following pirfenidone treatment(19). The findings of the current study align with previous research, underscoring the limitations of ^18^F-FDG in the early detection and therapeutic assessment of pneumoconiosis-induced pulmonary fibrosis. This highlights the superiority of ^18^F-FAPI for non-invasively evaluating active fibrotic lesions.

As a specific biomarker for detecting activated fibroblasts, ^18^F-FAPI can be utilized for real-time evaluation of pulmonary fibrosis activity in pneumoconiosis and monitoring the therapeutic response to anti-fibrotic drugs. Furthermore, since activated fibroblasts are the precursors of tissue fibrosis, ^18^F-FAPI PET/CT imaging can detect pulmonary fibrosis in pneumoconiosis earlier and more sensitively than other radioactive tracers. Consequently, this allows for an earlier diagnosis of active pulmonary fibrosis in pneumoconiosis patients, enabling timely treatment and potentially leading to a better prognosis. Additionally, ^18^F-FAPI PET/CT imaging may serve as a new non-invasive tool for the early detection of active fibrosis caused by various interstitial lung diseases. This can guide clinicians in choosing appropriate treatment timing, potentially reversing lung fibrosis to some extent, reducing lung structural damage, slowing the decline in lung function, and ultimately improving the prognosis of patients with pulmonary fibrosis.

The limitations of this study are as follows. First, the timing of treatment in the animal experiments began with dust exposure, whereas many pneumoconiosis patients in clinical practice started therapy after several years of dust exposure. Second, the clinical cases included in this study were limited to stage I pneumoconiosis with mild fibrosis, which introduces a potential selection bias. Moreover, the clinical study lacked an evaluation of treatment response and pathological verification.

In conclusion,^18^F-FAPI PET/CT imaging is a viable tool for the early detection and assessment of pulmonary fibrosis activity in pneumoconiosis, thereby facilitating timely and precise clinical interventions aimed at mitigating fibrotic progression. This modality exhibits superior sensitivity compared to ^18^F-FDG in monitoring the response to anti-fibrotic therapies, enhancing the precision of therapeutic adjustments based on treatment efficacy.

## Data Availability

All data produced in the present study are available upon reasonable request to the authors

## Abbreviations

CTGF: connective tissue growth factor
DL_CO_: diffusion lung carbon monoxide
ELISA: enzyme-linked immunosorbent assay
FAP: fibroblast activation protein
FAPI: fibroblast activation protein inhibitors
FVC: forced vital capacity
HRCT: high-resolution computerized tomography
IF: immunofluorescence
IHC: immunohistochemistry
IL-6: interleukin 6
PET: Positron Emission Tomography
SUV: standardized uptake value
TBR: target-to-background ratio
TNF-□: tumor necrosis factor-alpha

