## Supplemental Materials for "^18^F-FAPI PET/CT Imaging in Pneumoconiosis: a new tool for early diagnosis and guiding treatment of pulmonary fibrosis"

**Supplemental methods**

**S1. Pathological staining**

To detect tissue fibrosis, rat lung tissue specimens were collected for hematoxylin and eosin (HE) and Masson trichrome staining each month. Dripped 4% paraformaldehyde into the lung, fixed it overnight in the fixative at room temperature, and embedded it in paraffin blocks. Then, prepared lung slices (5 μm), deparaffinize, and stain with hematoxylin and eosin (H&E) or Masson trichrome staining according to the manufacturer's instructions. Masson’s stain images were analyzed using ImageJ software (National Institutes of Health, Bethesda, MD). Relative collagen content (%) was calculated from collagen-positive blue area / total observed area ×100%.

**S2. Immunofluorescence Staining**

FAP expression levels in lung tissue were determined using immunofluorescence (IF) method. Fixed sliced tissue was sealed with 5% BSA (Solarbio, A8010, Beijing Solarbio Science & Technology Co., Ltd.) for 1h, incubated with primary antibody (FAP Rabbit pAb, ABclonal, A6349, ABclonal Biotechnology Co., Ltd.) at room temperature for 1 h, and then incubated with secondary antibody (F2765, ThermoFisher) for 50 min. The slices were placed in a medium with 4', 6-diamido-2-phenylindole (Powerful Biology, Wuhan Powerful Biology Technology Co., Ltd.). The fluorescence images were collected using a fluorescence microscope (Leica Dmi8). Image J software (National Institutes of Health) was used for image analysis. The mean fluorescence intensity of the FAP was calculated from immunofluorescence-stained slides as fluorescence area (Intden) / total observed area (Area) ×100%.

**S3. Immunohistochemistry (IHC)**

Paraffin-embedded lung tissue sections (5 μm) were deparaffinized and rehydrated with graded ethanol, followed by antigen retrieval by microwave heating in citric acid at pH 6.0. Incubate with 3% H_2_O_2_ solution for 25 minutes to block endogenous peroxidase. After incubating with 3% BSA for 30 minutes, the rabbit anti-rat monoclonal antibody against FAP (FAP Rabbit pAb, ABclonal, A6349, ABclonal Biotechnology Co., Ltd.) was incubated overnight at 4 ℃ at a dilution of 1:50 and then incubated with biotinylated goat anti-rabbit IgG (ab205718, Abcam) at a dilution of 1:200 at room temperature for 50 minutes. All sections were stained with peroxidase-conjugated avidin/biotin and 39-3-diaminobenzidine (DAB) substrate (K3468, DAKO), followed by counterstaining with hematoxylin. Image analysis was performed using ImageJ software (National Institutes of Health, Bethesda, MD). FAP+ area% was calculated from IHC slides as Intden / Area ×100%.

**S4. Real-time Quantitative PCR (qPCR)**

Total lung tissue RNA was extracted using Trizol (TIANGEN, DP424, TIANGEN BIOTECH CO., LTD.). cDNA was synthesized using the Evo M-MLV Reverse Transcription Kit (AG11728, Accurate Biology). QPCR was conducted on QuantStudio 7 Flex (ABI). The primer sequence used were as follows: FAP: TCCACGGAACAGGAT (forward); GGTCAGAGTACCACATCGCC (reverse); GAPDH: TGACATCAAGAAGGTGGTGAAGC (forward), GGAAGAATGGGAGTTGCTGTTG (reverse). qPCR data represented target genes' relative mRNA expression levels, and GAPDH was used as an internal reference control.

**S5. Enzyme-linked immunosorbent assay**

Blood specimens were taken from rats at each time point from the abdominal aorta and then centrifuged in a vacuum tube at 3500 rpm for 10 min to obtain the plasma. Concentrations of plasma interleukin 6 (IL-6), tumor necrosis factor ɑ (TNF-ɑ), and connective tissue growth factor (CTGF) were measured using enzyme immunoassay kits (JIUBANG, ZC-10049, ZC-10160, ZC-10165, Quanzhou Jiubang Biotechnology Co., Ltd.).

**Supplemental figures**

**FIGURE** **S1.** Quantitative analysis of inflammation in rat lungs in each month.

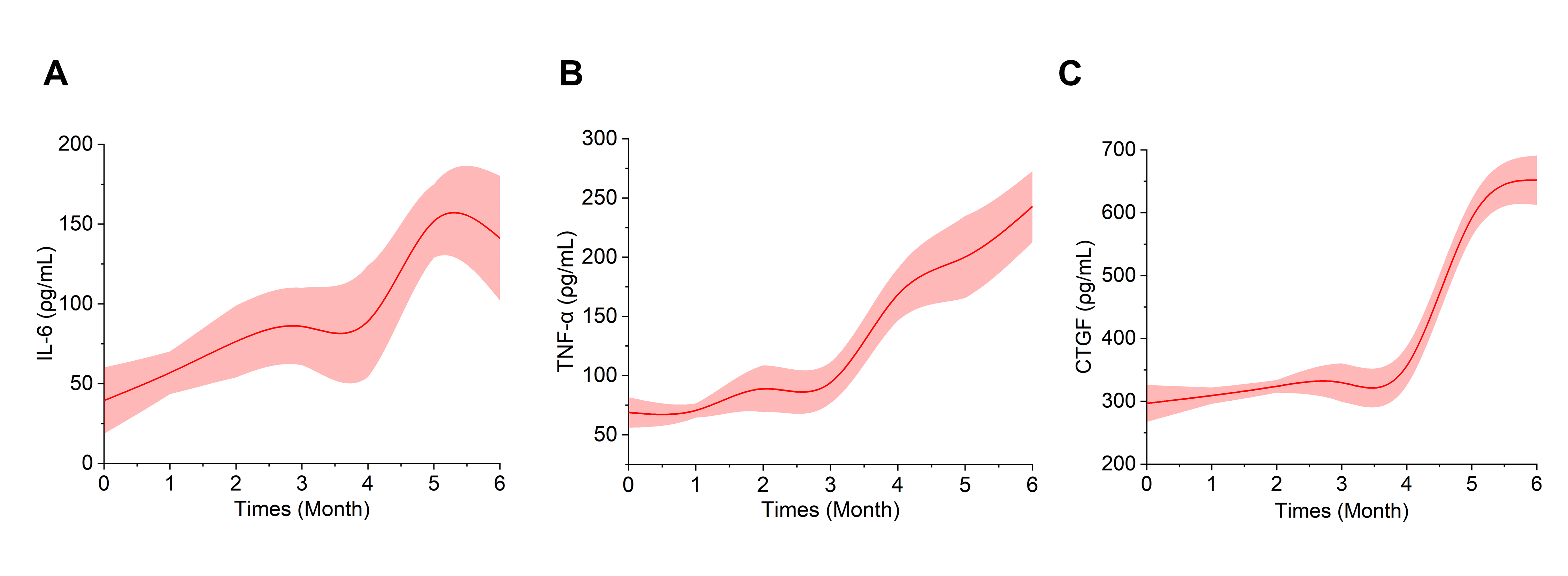

Caption: (d) Plasma levels of Interleukin 6 (IL-6) in the pneumoconiosis rat. (e) Plasma levels of tumor necrosis factor-alpha (TNF-ɑ) in the pneumoconiosis rat. (f) Plasma levels of connective tissue growth factor (CTGF) in the pneumoconiosis rat.
